# Multicenter point-prevalence evaluation of the utilization and safety of drug therapies for COVID-19

**DOI:** 10.1101/2020.06.03.20121558

**Authors:** Nathaniel J. Rhodes, Atheer Dairem, William J. Moore, Anooj Shah, Michael J. Postelnick, Melissa E. Badowski, Sarah M. Michienzi, Jaime L. Borkowski, Radhika S. Polisetty, Karen Fong, Emily S. Spivak, James R. Beardsley, Cory M. Hale, Andrea M. Pallotta, Pavithra Srinivas, Lucas T. Schulz, on behalf of the Vizient AMC Pharmacy Network Research and Antimicrobial Stewardship Committees

## Abstract

**Background:** There are currently no FDA-approved medications for the treatment of COVID-19. At the onset of the pandemic, off-label medication use was supported by limited or no clinical data. We sought to characterize experimental COVID-19 therapies and identify safety signals during this period.

**Methods:** We conducted a non-interventional, multicenter, point prevalence study of patients hospitalized with suspected/confirmed COVID-19. Clinical and treatment characteristics within a 24-hour window were evaluated in a random sample of up to 30 patients per site. The primary objective was to describe COVID-19 targeted therapies. The secondary objective was to describe adverse drug reactions (ADRs).

**Results:** A total of 352 patients from 15 US hospitals were included. Most patients were treated at academic medical centers (53.4%) or community hospitals (42.6%). Sixty-seven patients (19%) were receiving drug therapy in addition to supportive care. Drug therapies included hydroxychloroquine (69%), remdesivir (10%), and interleukin-6 inhibitors (9%). Five patients (7.5%) were receiving combination therapy. Patients with a history of asthma (14.9% vs. 7%, p = 0.037) and those enrolled in clinical trials (26.9% vs.

3.2%, p< 0.001) were more likely to receive therapy. Among those receiving COVID-19 therapy, eight patients (12%) experienced an ADR, and ADRs were more commonly recognized in patients enrolled in clinical trials (62.5% vs 22%, OR = 5.9, p = 0.028).

**Conclusions:** While we observed high rates of supportive care for patients with COVID-19, we also found that ADRs were common among patients receiving drug therapy including in clinical trials. Comprehensive systems are needed to identify and mitigate ADRs associated with experimental COVID-19 therapies.

## Introduction

Severe acute respiratory syndrome coronavirus 2 (SARS-CoV-2) is the pathogen responsible for the Coronavirus Disease 2019 (COVID-19) that has caused a global pandemic.^1^ There are a few potential therapies with activity against SARS-CoV-2;^2^ yet, no agent has received Food and Drug Administration (FDA) approval to treat COVID-19 to date. Investigational agents proposed at the onset of the pandemic included antimicrobials, remdesivir, lopinavir/ritonavir, nitazoxanide, ivermectin, and azithromycin. Host cell modulators including hydroxychloroquine, chloroquine, and agents targeting the host immune system have also been proposed.^2^ Use of any of these agents is expected to vary across care settings and over time as new clinical evidence and safety information becomes available.

Many of the investigational agents lack robust evidence to support their safety in COVID-19, which may increase the potential risk of undue harm. Each of these agents are associated with adverse drug reactions (ADRs) based upon prior research, yet little is known about the safety of these agents for use in patients with COVID-19.^3^ The safety and efficacy of investigational agents being used currently for patients with COVID-19 are only beginning to emerge.^4^ Safety concerns range from arrhythmias and QTc prolongation with hydroxychloroquine and azithromycin^5^ to intestinal perforation with tocilizumab.^6^ The lack of robust evidence on the safety of COVID-19 therapies has prompted the FDA to require clinicians to report serious adverse events associated with Emergency Use Authorization (EUA) remdesivir.^7^ A clear need exists for comprehensive data on ADRs with drug therapies targeting COVID-19.

ADRs associated with investigational and unproven therapies targeting COVID-19 may be difficult to detect and may vary with the usage rates of supportive care. Routine monitoring for potential ADRs is encouraged by consensus guidelines, particularly as use of investigational and EUA agents continues outside of rigorously monitored clinical trials.^4^ The multicenter point-prevalence methodology may allow detection of ADR signals on a large scale and is relatively easy to implement rapidly. Although this methodology does not allow investigators to infer causation, it does supply valuable information about the landscape of current practice and may discern effects that otherwise could be missed within a single center.^8, 9^ We conducted a multicenter point-prevalence study to evaluate the drug therapies used to treat COVID-19. The objective of this point-prevalence study was to characterize the drug therapies used in the management of COVID-19, including supportive care and combination therapies, in an attempt to identify safety signals among acutely ill hospitalized patients.

## Patients and methods

### Study design

We conducted a non-interventional, multicenter, point-prevalence study of the medical records for hospitalized patients with COVID-19. Patients were identified at each site according to institutional guidelines for identification and monitoring of COVID-19 patients. The study was reviewed by each participating organization’s individual Institutional Review Board and found to be exempt. A waiver of informed consent and HIPAA authorization was completed at each site.

Patients were eligible for inclusion if they were hospitalized as inpatients and 1) had a positive SARS-CoV-2 test completed or 2) had a clinical diagnosis of COVID-19 based on a physician’s diagnosis. No limitations were placed on time from diagnosis to inclusion. We did not extract information on protected status with the exception of pediatric status; protected elements not evaluated in this study included dates of symptom onset or duration of hospitalization prior to evaluation. Patients were excluded if they were initially treated for COVID-19, but an alternative diagnosis was made prior to being evaluated for the survey (i.e., COVID-19 was ruled-out).

### Data elements

The point-prevalence survey was circulated on April 18, 2020 to 15 hospitals across the United States (US). Each site was asked to select a random sample of up to 30 patients and to complete data collection within 1 month. Any hospitalized patient meeting inclusion criteria was eligible. Data were manually extracted from the electronic health records and was entered into a standardized electronic survey form. Study data were collected and managed using REDCap electronic data capture tools hosted at Northwestern University. REDCap (Research Electronic Data Capture) is a secure, web-based software platform designed to support data capture for research studies.^10, 11^ Data validation was coordinated by the Principal Investigator and each respective site coordinator. Data elements collected included facility demographics, total number of hospital and ICU beds prior to the pandemic, U.S. Census region location, patient populations served, facility type (e.g., academic, community, inpatient rehabilitation), and active clinical trial site status. Given the non-interventional nature of this study, all patients were managed at each site according to each center’s standard of care and guidelines for the management of COVID-19.

In addition to whether patients were receiving supportive care or drug therapies targeted at SARS-CoV-2, we collected basic patient demographic information and vital status (e.g., age, sex, comorbidities, oxygen requirement, and ICU status). Data were abstracted from the electronic medical record – EMR (e.g., EPIC or Cerner depending on the EMR in use at the site) by infectious diseases and antimicrobial stewardship pharmacists at each site. For each patient included no follow-up or longitudinal outcomes were ascertained. Data extraction from the electronic health records were limited to the prior 24 hours, and no identifiers were collected. Pediatric patients were eligible for inclusion.

### Study definitions

For patients aged > 90 years, age was classified as > 90 years. We evaluated the use of the following specific medications: azithromycin (AZM), hydroxychloroquine (HCQ), tocilizumab or sarilumab (IL-6 inhibitors), lopinavir/ritonavir (L/R), remdesivir (clinical trial or compassionate use), or other investigational agents. Clinical trial enrollment status was evaluated based upon notes within the medical record. Combination therapy was considered as concurrent receipt of more than one agent targeting SARS-CoV-2 (e.g., HCQ+AZM or HCQ+remdesivir). Requirement and degree of oxygen support within the last 24 hours was collected (i.e., no oxygen needed, supplemental oxygen required, mechanical ventilation; low flow, high flow, or invasive ventilation). ICU admission status was also recorded.

Adverse drug reactions were defined based upon existing knowledge of the side effect profiles of the agents evaluated as well as the existing literature on adverse events observed in COVID-19 patients at the time of the study. Attribution of ADRs were based upon the electronic health records and upon the clinical discretion of the data extractors. We evaluated the presence of new or worsening ADRs occurring concurrent to COVID-19 drug therapies and observed the following reactions within the prior 24 hours: transaminitis (i.e., both 3x and 5x the upper limit of normal), acute kidney injury (i.e., an increase in serum creatinine of 0.3 mg/L or 50%^12^), coagulopathy, headache, diarrhea, nausea or vomiting, prolonged QTc (i.e., > 500 ms), new onset arrhythmia, neutropenia, or thrombocytopenia. Worsening clinical status as documented within a physician’s note or in-hospital mortality were also evaluated as safety endpoints.

### Statistical analysis

Continuous data were analyzed using the Student’s t-test or Wilcoxon rank-sum, as appropriate. Categorical data were analyzed using Chi-square or Fisher’s Exact test, as appropriate. Missing data were treated as missing. All statistical analyses were performed using Intercooled Stata version 14.2 (College Station, TX). Statistical significance was set at alpha < 0.05. Univariate logistic regression was used to estimate OR for harm only when the event rate exceeded 10% for the outcome (e.g., any ADR).

## Results

### Demographics

A total of 352 patients admitted to 15 hospitals across the US between April 18 and May 8, 2020 were included. Patient and facility demographics are summarized and stratified according to receipt of only supportive care vs COVID-19 directed therapy in **Table 1**. Patients were primarily treated at academic medical centers (53.4%), followed by community hospitals (42.6%), and a minority at rehabilitation hospitals (4%). The majority of patients in our study were treated in the Midwest US region (81%). Over 97% of patients were confirmed to have COVID-19 based on diagnostic testing, with 98% of specimens collected via nasopharyngeal swab and minority collected via bronchoalveolar lavage. The mean [standard deviation (SD)] age of patients was 61.9 (16.1) years. Fifty-two percent of patients were males. The mean (SD) body weight and BMI were 89 (29) kg and 31.8 (11.3) kg/m^2^, respectively. Over 95% of patients were adults but 4.5% were pediatric or neonatal patients. The majority of patients (81%) were receiving supportive care only (including supplemental oxygen and/or other non-pharmacological support).

**Table 1.**
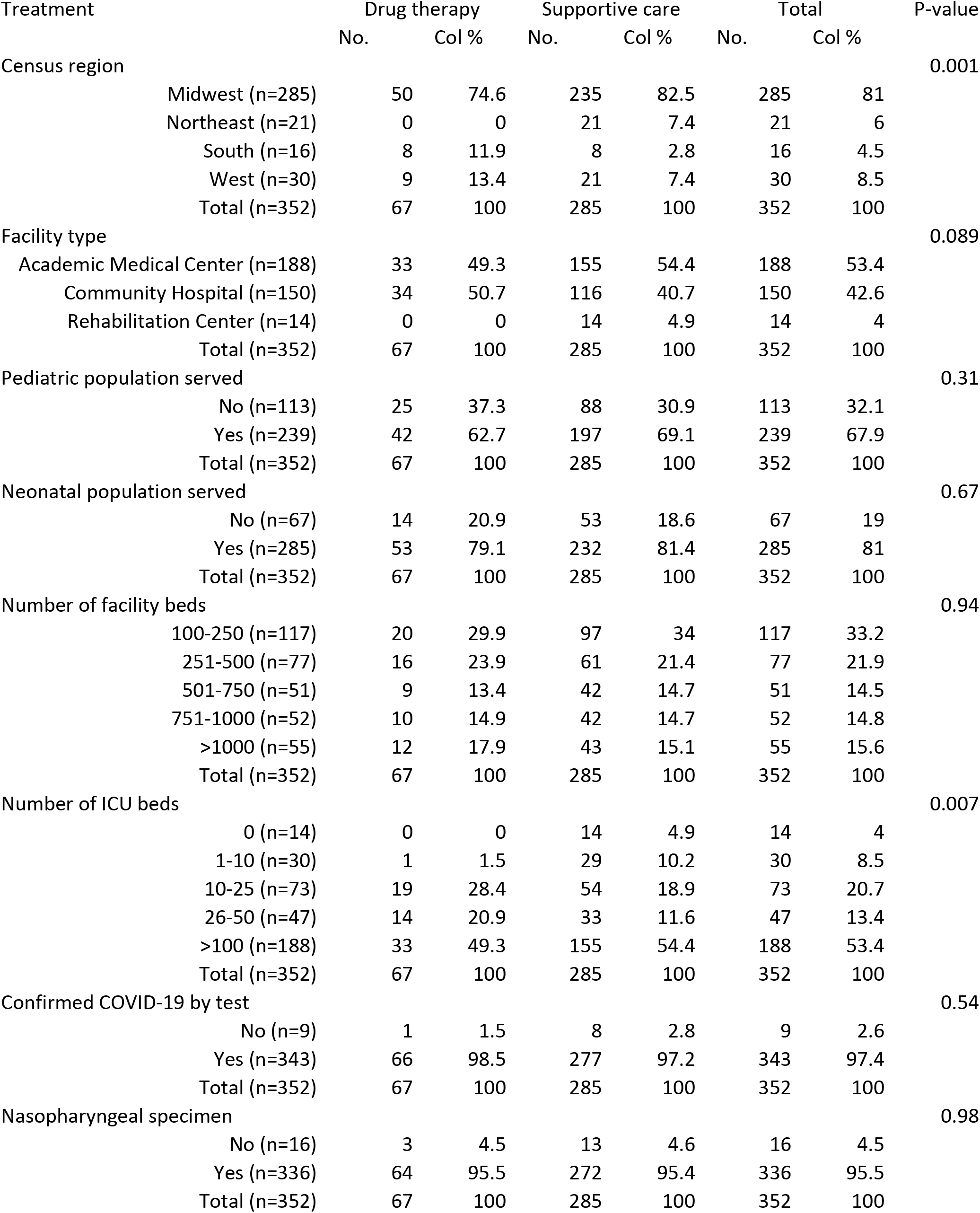
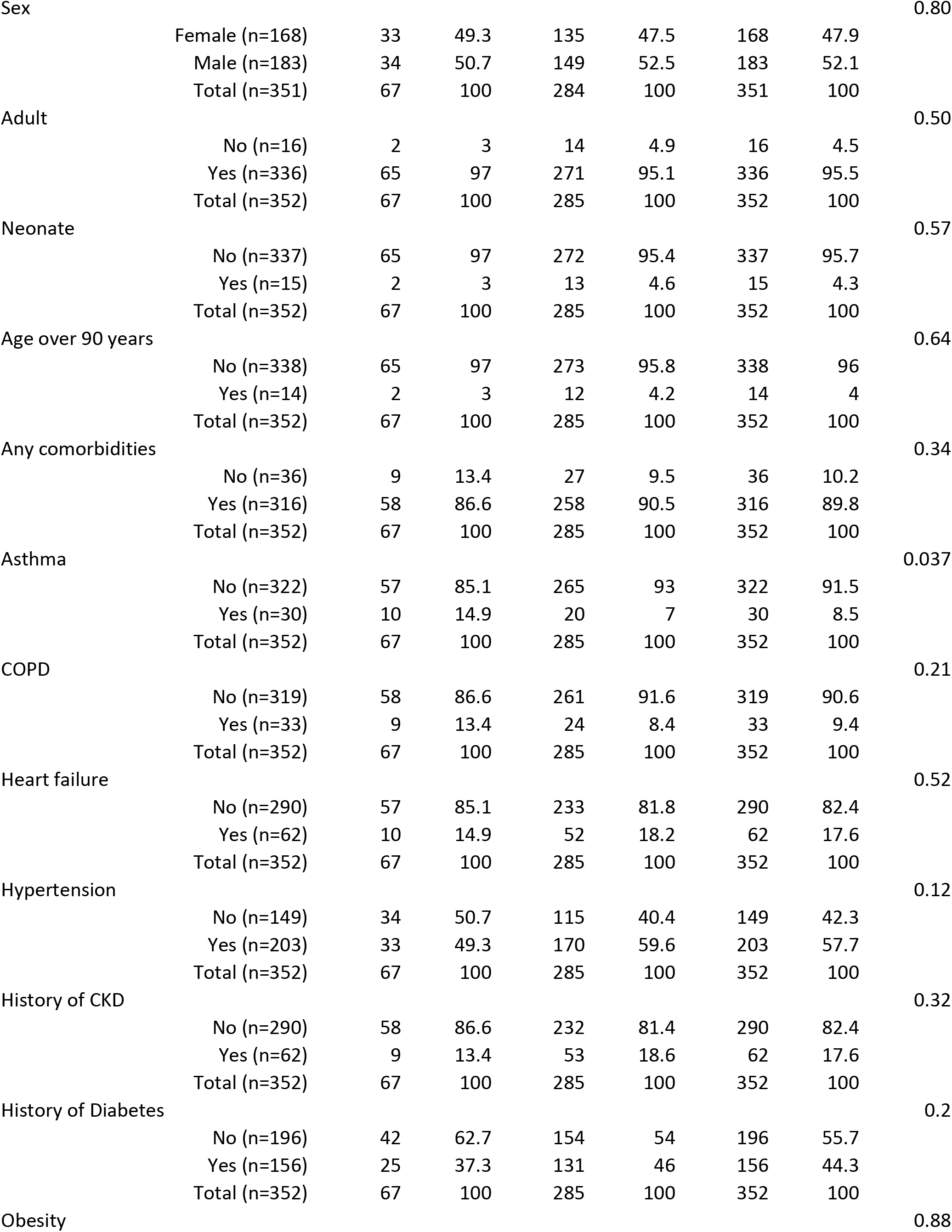
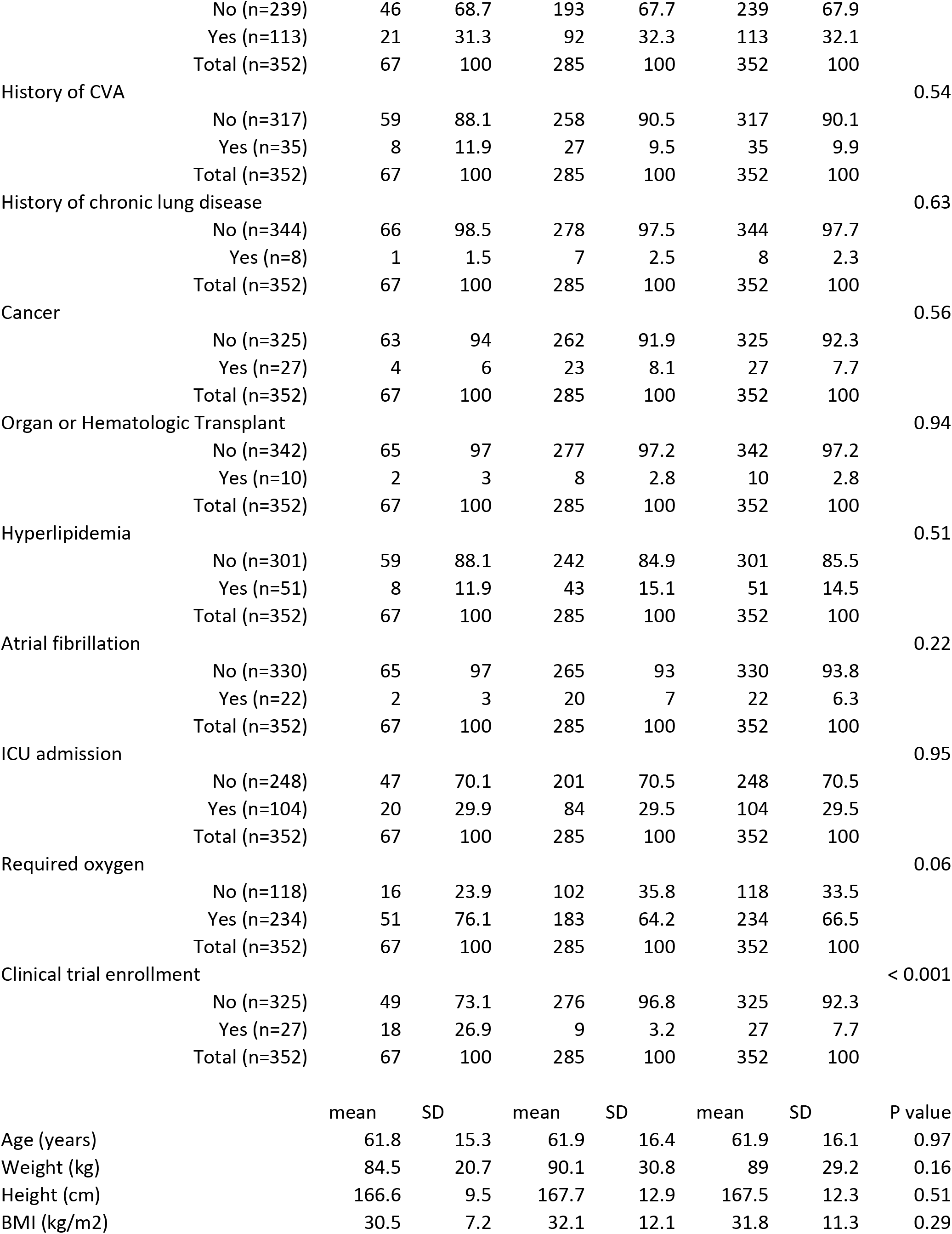
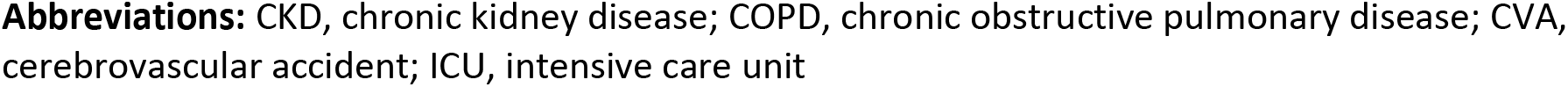
Patient and facility demographics according to the use of only supportive care or COVID-19 directed therapy

### Supportive care vs COVID-19 directed therapy

A total of 67 patients were receiving COVID-19 directed drug therapy. The most common COVID-19 directed drug therapies used were HCQ (69%), remdesivir (10%), and IL-6 inhibitors (9%). A total of 51 of the 67 patients (76%) treated with COVID-19 directed therapies required supplemental oxygen. Patients with a history of asthma were significantly more likely to receive COVID-19 drug therapy vs supportive care only (14.9% vs 7%; P = 0.037). Patients were significantly more likely to receive COVID-19 directed drug therapy if they were enrolled in a clinical trial (26.9% vs 3.2%; P < 0.001).

### Frequency of ADRs in patients receiving monotherapy vs combination COVID-19 directed therapy

Among patients receiving COVID-19 therapy, a total of 8 patients (12%) experienced any ADR, five of which (7.5%) were receiving combination treatment. A summary of the types and frequency of ADRs with combination and monotherapy COVID-19 directed therapy are summarized in **Table 2**. All 5 of the patients who were receiving combination therapy were given HCQ in addition to another agent directed at SARS-CoV-2 including: azithromycin (n = 2/5), an IL-6 blocker (n = 2/5), or some other investigational agent (n = 1/5). Patients who were receiving combination therapy were numerically more likely to experience any ADR (20% vs 11.3%; P = 0.56). Patients who were receiving monotherapy and combination therapy did not differ with respect to any specific ADR evaluated (**Table 2**), but numerically more patients experienced diarrhea if they were given combination therapy (20% vs 4.8%; P = 0.27). Patients with a history of chronic kidney disease (CKD) were numerically more likely to have any ADR detected (37.5% vs 10.2%; P = 0.068). Likewise, patients enrolled in any clinical trial were significantly more likely to have any ADR detected compared to patients who were not enrolled in a clinical trial (62.5% vs 22%; OR = 5.9; P = 0.028).

**Table 2.** Frequency and type of adverse drug reactions according to combination COVID-19 directed therapy

|  | Monotherapy |  | Combination therapy |  | Total |  | P-value |
| --- | --- | --- | --- | --- | --- | --- | --- |
|  | No. | Col % | No. | Col % | No. | Col % |  |
| Any ADR in last 24 hr |  |  |  |  |  |  | 0.48 |
| No (n=59) | 55 | 88.7 | 4 | 80 | 59 | 88.1 |  |
| Yes (n=8) | 7 | 11.3 | 1 | 20 | 8 | 11.9 |  |
| Total (n=67) | 62 | 100 | 5 | 100 | 67 | 100 |  |
| AKI |  |  |  |  |  |  | >0.99 |
| No (n=65) | 60 | 96.8 | 5 | 100 | 65 | 97 |  |
| Yes (n=2) | 2 | 3.2 | 0 | 0 | 2 | 3 |  |
| Total (n=67) | 62 | 100 | 5 | 100 | 67 | 100 |  |
| Diarrhea |  |  |  |  |  |  | 0.27 |
| No (n=63) | 59 | 95.2 | 4 | 80 | 63 | 94 |  |
| Yes (n=4) | 3 | 4.8 | 1 | 20 | 4 | 6 |  |
| Total (n=67) | 62 | 100 | 5 | 100 | 67 | 100 |  |
| Prolonged QTc > 500 ms |  |  |  |  |  |  | >0.99 |
| No (n=65) | 60 | 96.8 | 5 | 100 | 65 | 97 |  |
| Yes (n=2) | 2 | 3.2 | 0 | 0 | 2 | 3 |  |
| Total (n=67) | 62 | 100 | 5 | 100 | 67 | 100 |  |
| Thrombocytopenia |  |  |  |  |  |  | >0.99 |
| No (n=64) | 59 | 95.2 | 5 | 100 | 64 | 95.5 |  |
| Yes (n=3) | 3 | 4.8 | 0 | 0 | 3 | 4.5 |  |
| Total (n=67) | 62 | 100 | 5 | 100 | 67 | 100 |  |
| ADR led to discontinuation |  |  |  |  |  |  | >0.99 |
| No (n=7) | 6 | 85.7 | 1 | 100 | 7 | 87.5 |  |
| Yes (n=1) | 1 | 14.3 | 0 | 0 | 1 | 12.5 |  |
| Total (n=8) | 7 | 100 | 1 | 100 | 8 | 100 |  |
| Worsened in last 24 hr |  |  |  |  |  |  | >0.99 |
| No (n=51) | 47 | 75.8 | 4 | 80 | 51 | 76.1 |  |
| Yes (n=16) | 15 | 24.2 | 1 | 20 | 16 | 23.9 |  |
| Total (n=67) | 62 | 100 | 5 | 100 | 67 | 100 |  |
| Mortality in last 24 hours |  |  |  |  |  |  | >0.99 |
| No (n=66) | 61 | 98.4 | 5 | 100 | 66 | 98.5 |  |
| Yes (n=1) | 1 | 1.6 | 0 | 0 | 1 | 1.5 |  |
| Total (n=67) | 62 | 100 | 5 | 100 | 67 | 100 |  |

**Abbreviations:** ADR, adverse drug reaction; AKI, acute kidney injury

### Frequency of COVID-19 directed therapy and ADRs among patients requiring oxygen therapy

The requirement for supplemental oxygen secondary to COVID-19 was numerically more common among patients who were receiving combination COVID-19 directed therapy compared to monotherapy [77.4% (n = 48/62) vs 60% (n = 3/5); P = 0.59). A summary of adverse effects according to oxygen requirement status is summarized in **Table 3**. Patients who did not require oxygen were numerically more likely to experience any ADR within the prior 24 hr (25% vs 7.8%; P = 0.085). These patients were also numerically more likely to experienced QTc prolongation within the prior 24 hr (12.5% vs 0%; P = 0.054). On the other hand, patients who required supplemental oxygen were significantly more likely to have clinically worsened in the prior 24 hours (31.4% vs 0%; P = 0.008). We did not observe significantly more death among patients who required supplemental oxygen (P = 0.57).

**Table 3.**
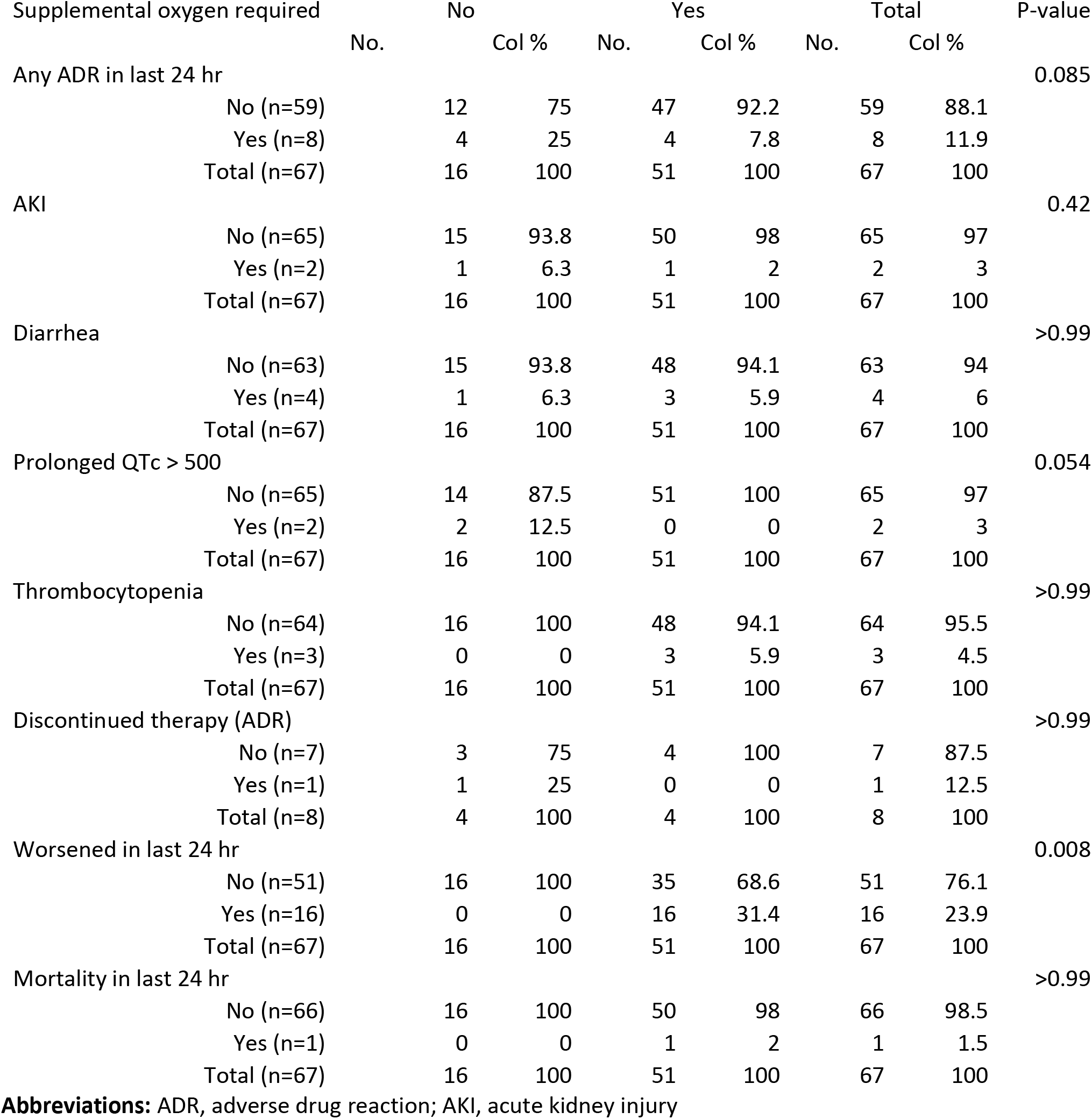
Frequency of adverse drug reactions according to supplemental oxygen requirement

## Discussion

This multicenter point-prevalence study found that, between 4/12/20-5/8/20, drug therapy for COVID-19 was relatively uncommon across academic, community, and rehabilitation hospitals during a typical day. The majority of patients were receiving supportive care (81%) and a total of 66.5% required supplemental oxygen. Only 67 patients (19%) were receiving COVID-19 directed drug therapy, and a minority of patients were receiving combination treatment with more than one agent targeting SARS-CoV-2 (n = 5). Nevertheless, we still were able to identify ADRs in 12% of patients who were receiving drug therapy. While a point-prevalence approach is not comprehensive and causality cannot be firmly established, our findings serve as a warning that COVID-19 drug therapies are not completely benign. Among patients receiving combination therapy (mostly involving HCQ) diarrhea was more common. Our findings suggest that, on a typical day, more patients were receiving supportive care compared to COVID-19 drug therapies. Enrollment in a clinical trial and a history of asthma were associated with increased use of drug therapies targeting SARS-CoV-2.

Notably, our sample represents 15 hospitals distributed throughout the United States, with high representation from the Midwest; a region underrepresented in the available COVID-19 literature. It is also important to note that our study took place before wide availability of EUA remdesivir.^13^ Most patients in our study were receiving supportive care at the time of evaluation; however the cumulative exposure to drug therapies was not ascertained. Supportive care has been suggested as a cautious approach to providing unproven and understudied drug therapy in an emergency situation,^14, 15^ and more data are needed to define the safety of COVID-19 drug therapies. Our finding that ADRs were relatively common among COVID-19 patients receiving drug-therapy underscores the need for close monitoring. We found that patients enrolled in clinical trials were nearly 6-fold more likely to have an ADR detected, likely reflecting the careful monitoring occurring in trials. Unfortunately, the vast majority of patients with COVID-19 do not have access to clinical trials but receive these agents, nonetheless. Therefore, a clear need exists for improved ADR monitoring in patients receiving unproven therapies.

As noted, various medications are being evaluated in the fight against COVID-19; however, at the time of this writing there is no definitive cure. While some drugs have been touted as efficacious, clinical trial findings have not mirrored these claims and many of these potential treatments have been plagued by safety concerns. Lopinavir/ritonavir was initially evaluated in China for its role in hospitalized adults diagnosed with severe COVID-19, but it failed to demonstrate benefits including time to clinical improvement or mortality at 28 days when compared to standard of care.^16^ Serious adverse events were more common in the standard care group yet lopinavir/ritonavir was associated with more gastrointestinal ADRs. Of note, we did not observe any use of lopinavir/ritonavir in our patient sample.

Similarly, the use of HCQ, alone or combined with azithromycin, has diminished markedly due to safety concerns and questionable efficacy.^17, 18^ Though less than 20% of patients in our study were receiving COVID-19 drug therapy, the most commonly used drug was HCQ. Recently, results of a multinational registry analysis of over 96,000 hospitalized COVID-19 patients treated with HCQ (n = 3016), HCQ + macrolide (n = 6221), chloroquine (n = 1868), or chloroquine + macrolide (n = 3783) were reported.^18^ The investigators observed an increased risk of in-hospital mortality and de-novo ventricular arrhythmias with HCQ or chloroquine-based therapy, though these results have been called into question more recently.^19^ Subsequently, the World Health Organization paused enrollment for a clinical trial of HCQ, and the fate of HCQ as a COVID-19 therapy is currently uncertain.^20^

Whereas the agents above have known limitations, other agents including IL-6 inhibitors (tocilizumab, sarilumab), IL-1 inhibitors (anakinra, canakinumab), and remdesivir are gaining increased attention. The IL-6 inhibitor, tocilizumab, appeared beneficial in critically ill patients with severe COVID-19 based upon improvements in oxygen requirements, fever resolution, lung imaging, and inflammatory markers.^21^ Preliminary data from randomized, placebo-controlled, trial of sarilumab showed clinical improvements in patients with critical COVID-19.^22^ However, these treatments are not without risks as agents targeting the cytokine response may increase the risk for secondary infections, gastrointestinal perforations, and hepatic toxicity.^23^ Comprehensive information regarding the safety of these as yet unproven agents is needed as their use becomes more common in patients with COVID-19.

Remdesivir has garnered considerable interest as a treatment for COVID-19 since May 2020 when it was granted EUA status.^13^ A recent randomized placebo controlled trial by Wang et al. did not establish significant benefit of remdesivir and was terminated early.^24^ Serious adverse events occurred in 18% and 26% of patients receiving remdesivir and placebo, respectively. ACTT-1 was a randomized, placebo-controlled, study of 1,063 hospitalized patients with COVID-19.^25^ Preliminary results showed a shorter median time to recovery with remdesivir (11 vs 15 days). Serious adverse events occurred in 21% and 27% of patients receiving remdesivir and placebo, respectively. ADRs occurring more often with remdesivir were decreased renal function, pyrexia, and hyperglycemia.^25^ The Gilead-sponsored SIMPLE trial comparing 5 and 10 days of remdesivir did not identify a primary efficacy difference for the regimens evaluated; however, significantly more patients treated for 10 days experienced any serious adverse event including AKI.^26^ We did not identify higher rates of ADRs with remdesivir in our study, but patients could have been receiving remdesivir as compassionate use or may have received placebo in the setting of a clinical trial. Based upon available evidence, patients receiving remdesivir, particularly for more than 5 days, will require more intensive monitoring.

Our study has some limitations. First, this was a point-prevalence survey and cause and effects are not discerned. Our survey data window was limited to the prior 24 hours, so it is likely that our evaluation of adverse effects is conservative. Because our ability to discern causation was limited, it is not completely clear to what extent the adverse events we found were related to COVID-19 vs drug therapies. Nevertheless, we observed numerically more adverse effects among patients who did not require supplemental oxygen. These patients were at various points of their disease course (i.e., we included anyone who was hospitalized and required treatment), so prior exposure to agents that may have led to the noted adverse effects (e.g., diarrhea from antibiotics) was not comprehensively assessed. More work is needed to define the time course of ADRs in this population in order to improve patient safety. Despite these limitations, strengths of our study include the representative sample of patients with COVID-19 across various geographic and demographic populations. Data from clinical trials and ADR registries are needed to more clearly define the risks of COVID-19 drug therapies.

In conclusion, we observed high rates of supportive care in more than 80% of included patients with COVID-19 in our point-prevalence survey. Among patients who were receiving drug therapy, we found that as many as 12% experienced an ADR within the prior 24 hours with the most commonly reported adverse event being diarrhea. Patients enrolled in clinical trials were over 6-fold more likely to have an ADR identified, suggesting a greater need for ADR monitoring in patients with COVID-19. Comprehensive systems are needed to identify and mitigate adverse effects associated with COVID-19 drug therapies.

## Data Availability

Data will be made available upon reasonable request from the authors. Such requests will be considered by a committee consisting of the investigators. Data will be made available pursuant to appropriate data use agreements.

## Acknowledgement

The Vizient Research Committee would like to acknowledge the following individuals for their assistance with data collection: Christie Bertram, PharmD, Stephanie Chang, PharmD, Ara Gharabagi, PharmD, Kirsten Robles, PharmD, Brian Hoff, PharmD, Elizabeth Neuner, PharmD, Maryam Rauf, PharmD, Rishita Shah, PharmD, and Sheila Wang, PharmD

